# Prevalence, associated factors and antimicrobial susceptibility patterns of *Salmonella* species and pathogenic *Escherichia coli* isolated from broiler poultry farms in Wakiso district, Uganda

**DOI:** 10.1101/2024.08.16.24312101

**Authors:** Thomas Ssemakadde, Nalumaga Pauline Petra, Jude Collins Busingye, Joel Bazira, Kabanda Taseera

## Abstract

**Background:** The emergence and re- emergence of zoonotic bacterial infections and the upsurge reflected in current trends of antimicrobial-resistant bacteria is a major global concern. *Salmonella* spp and *Escherichia coli (E. coli)* are the two most important food-borne pathogens of public health interest incriminated in poultry products worldwide hence necessitating constant monitoring of microbial food safety measures. The purpose of this study was to determine the prevalence, associated factors and antimicrobial susceptibility patterns of *Salmonella* and *E. coli* in poultry farms in Wakiso District to provide detailed information of extent of spread to guide plans that influence safer poultry keeping practices in this era.

**Methods:** This study was a cross sectional study that used a total of two hundred sixteen(216) poultry samples from cloacae swabs and fecal swabs collected from broiler poultry farms and cultured on Chromagar ^TM^ Salmonella and Sorbitol MacConkey agar for pathogenic E. coli. Biochemical tests, minimum inhibitory concentration, and polymerase chain reaction were utilized. Assessment of the correlations between the resistance patterns of resistant and susceptible isolates was determined using mean, and multiple logistic regression.

**Results:** A total of 40 (18.5%) *Salmonella* and 120 (55.6%) Pathogenic *E. coli* was isolated. In this investigation, extended beta lactamase (ESBL) production was detected in 18 isolates *Salmonella* and 57 pathogenic *E. coli*. Prevalence of *bla*TEM gene was expressed in 7/18 (39%) *Salmonella* isolates and 42/57 (73.8%) Pathogenic *E. coli* isolates The associated factors that predispose these farms to *Salmonella* and Pathogenic *E. coli* identified in this study include: contact of poultry and wild birds (p- value =0.000), movement from one pen to the other by farm-handlers (P-Value = 0.030), use of untreated water (P-Value =0.005) and food contamination of commercial poultry feeds (P-Value= 0.0021)

**Conclusion:** *Salmonella* spp and *Escherichia coli* remain the two most important food-borne pathogens of public health interest incriminated in the poultry field, and it is evident from this study that these bacteria have resistant genes associated with them.

## INTRODUCTION

The emergence and re-emergence of zoonotic bacterial infections and the upsurge reflected in current global trends of antimicrobial resistance is a major universal concern (1). Presence of bacteria that have resistant genes decreases the efficacy of the few available antibiotics culminating into infection and failure patterns in treatment which sometimes are implicated in morbidity and mortality trends across the globe (2).

Globally poultry farming is one of the fast-growing low-cost investment ventures currently on the rise and this is partly explained by the significant growth in the population size that provides ready market for poultry products (3). As a result, poultry raised in rigorous, intense circumstances using antimicrobial drugs to hasten growth and prevent disease as a result of the method of raising the birds (4). Antibiotic-resistant bacteria in poultry could have resistant genes that could potentially be passed on to humans posing threat such as economic losses in animal production and treatment failure, leading to mortalities and morbidities (5).

According to WHO nearly 33 million healthy years of life worldwide are lost each year as a result of unsafe practices related to food handling leading to consumption of unsafe food (6). In Africa today, over 91 million people fall ill due to food borne illnesses and approximately 137,000 deaths are registered representing a third (1/3) of the global death toll from food borne illnesses. Diarrheal diseases in the African continent are estimated to account for 70% of the burden of food borne diseases (6).

Several studies done focusing on the disease trend patterns have highlighted the significant role of *E.coli* as a contributing member to the diarrheal disease burden in humans and this accounts for 4.0% of the global burden of disease daily thus contributing to an estimated 1.8 million fatalities each year, of which children account for roughly 90% (7, 8). In the poultry industry, Avian pathogenic Escherichia coli (APEC) is a typical bacterium that is proportionally linked to tremendous economic losses that is due to a number of conditions which range from septicemia, salpingitis, chronic respiratory disease, and embrionary death. For the recent colibacillosis outbreaks across the globe that represent 80% of the disease cases worldwide, the strongly associated serotypes are O78 and O2 (9). According to disease surveillance and outbreak data published in the MOH weekly Epidemiological bulletin, there is an upsurge in the number of Typhoid cases seen in Kampala and Wakiso districts (10). Owing to the upward increase in the human population, the demand driven venture of poultry rearing and the increase in typhoid cases, there is need to investigate whether these poultry products play a part in perpetuating these infections. The existence of these food borne pathogens in poultry leads to negative effects on both humans, animal, and environmental health sectors and as such calls for investigations to draft better methods of eliminating potential AMR situations.

This study will determine the antimicrobial susceptibility patterns, related variables, and prevalence of *Salmonella* and pathogenic *E. coli* in poultry farms in Wakiso District to provide detailed information of extent of spread, resistant genes and this will guide plans to influence safer poultry keeping practices in this era of increasing population pressure for poultry products.

## MATERIALS AND METHODS

### Study Area

This study was conducted in Wakiso district (as seen in figure 1) a metropolitan district in the central region of Uganda that partly encircles Kampala district. The district coordinates are: 00 24N, 32 29E. With a projected population of 2,915,200 million, Wakiso district is still Uganda’s most populous Higher Local Government (HLG) (11).There is an annual population increase projected at 4.1% and this rapidly increasing population increases the demand of food including poultry products. The district was regarded as the top producer of poultry, producing 7.4% of all the chickens in the country (12).

**Figure 1;.**
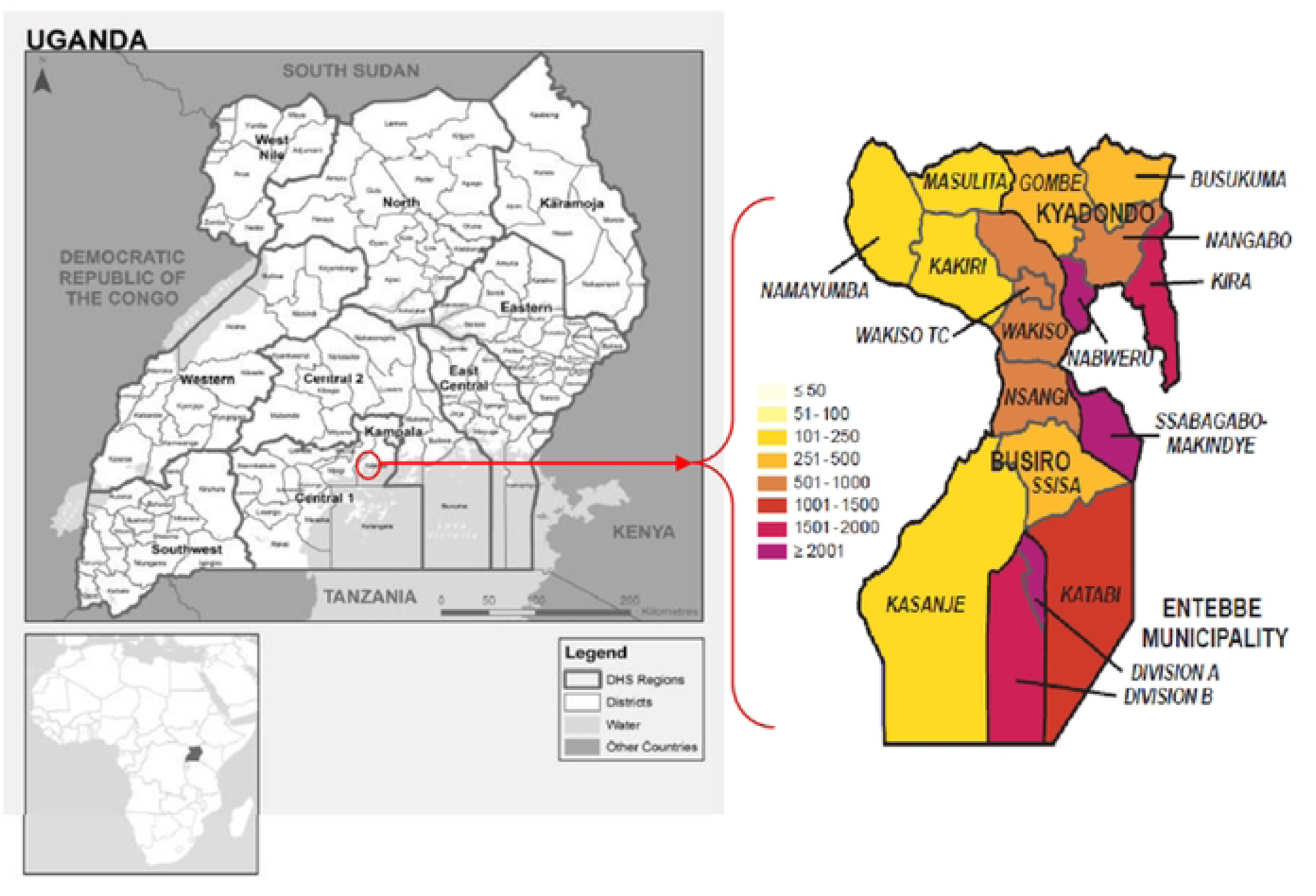
Highlighting Wakiso district and *its position on the Map of Uganda*.

### Study design

This was a cross-sectional study focused on broiler farms based on the FAO categorization sector level 3 farms whose chicken that was about to enter the food supply chain, within selected counties in Wakiso district. Fresh fecal droppings plus cloacal samples were collected during August to September period of the year 2021.

### Sample Size Determination

The sample size was determined using Kish-Leslie’s (1965) standard formula. The sample size was derived using 83% prevalence (13).

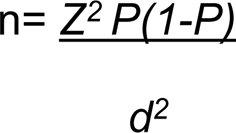

n = study sample size required

Z= critical value associated with 95% confidence interval = 1.96

P = Estimated prevalence of 83%of *E coli* and salmonella d = margin of error = 0.05

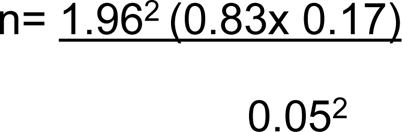

n= 216 samples

Therefore, a minimum of 216 samples were collected from the various farms in the different constituencies of Wakiso District. A subsequent number of farm managers with consent from these 216 farms were interviewed to understand the associated factors that predispose these pathogens to the farm.

### Sampling method

The study employed a variety of methods including simple random sampling to select the sub counties, purposive sampling using information provided by the district veterinary office to identify the villages and the respective farms from which samples were picked.

For the selected and eligible farms, three swabs from fresh fecal droppings and two swabs from the cloaca were collected while adhering to the appropriate bio safety measures at the farm as stated in the WHO bio safety manual fourth Edition. Five swabs from one farm on different chicken were considered to represent one sample (Pooling of samples).

### Sample Collection

Three fresh fecal droppings of chicken from the target shed with oldest flock were randomly selected. Using a sterile swab, adequate fecal material was obtained by swabbing the top of the freshly deposited fecal matter.

Two random chickens from the target shed with oldest flock were randomly selected. Using a sterile swab, adequate fecal material was obtained from the cloaca. The swabs were placed into a plain tube containing Amies transport media without charcoal. A Stopper was used to seal the tube, the sample containing tube was labeled with the sample identifier/ Farm research generated IDs, location and date and stored in a rack or zip lock bag. This was then put into a cool box and transported to the laboratory within 24 hours.

### Isolation and identification of microorganisms

#### For Pathogenic E. coli and Salmonella spp

The pooled samples were aseptically mixed into 9 ml of autoclaved Buffered Peptone Water (BPW) (Hi Media M1494, Mubi, India) in a sterile 50ml falcon tube with a lid to generate a pre-enriched sample, vortex, and incubated for 16-24 hours at 37°C aerobically.

The enriched sample was picked and inoculated onto a chromogenic agar (Chromagar^TM^ Salmonella which contains Agar, Peptone and yeast extract +Chromagenic& selective mix) which is a particular chromogenic culture medium designed for the presumptive identification, qualitative direct detection, and differentiation of Salmonella (14). Incubation of the plates was done for 24hrs at 37^0^C. Presumptive colonies for Salmonella (mauve-pink, raised and smooth colonies) were selected for identification

For isolation of pathogenic *E.coli*, the pre-enriched sample was inoculated on Sorbitol MacConkey agar plates and incubated at 37 °C ± 1°C for 24 hours. The colonies of pathogenic E. coli were smooth, raised, had entire margins and were colorless on SMAC. The presumptive organisms were subcultured on nutrient agar and identified through gram staining, Triple Ion Sugar (TSI) test, the IMViC test, Methyl Red, Voges-Proskauer, Sulphur indole motility, and Simmons citrate utilization. All reagents used were from (Oxoid, England) (15).

### Bacterial antibiotic susceptibility testing

Bacterial suspension that was adjusted to 0.5McFarand standard and inoculated on Muller Hinton agar medium (Oxoid CM0337 Basingstoke, England) using surface spreading method. The antibiotic discs listed in table 1 (all from Oxoid, England) were used. The plates were incubated for 24 hours at 37°C. The results were read and interpreted according to Clinical Laboratory Standards Institute standards Version 14th edition, 2020 (16).

**Table 1:** Commonly used antibiotics at NaLIRRI, 2022 obtained from Oxoid ltd suppliers and the discs were selected based on CLSI (28^th^ Edition, 2020) and their availability at NALIRRI Microbiology laboratory.

| Antibiotic Class | Antibiotic used for both *E. coli* and *Salmonella* isolates |
| --- | --- |
| Aminoglycosides | Gentamicin (GM) (10 µg/ml) |
| Carbapenem | Meropenem (MEM) (10 µg/ml) |
| Cephalosporins class III | Ceftriaxone (CRO) (30 µg/ml) |
| Amphenicol | Chloramphenicol (CHL) (30 µg/ml) |
| Cephalosporins class IV | Cefepime (CPM) (30 µg/ml), Cefotaxime (CTX)(30µg/ml), Ceftazidime (CTZ) (30µg/ml) |
| Quinolones | Ciprofloxacin (CIP) (5 µg/ml) |
| Macrolides | Erythromycin (EM) (30 µg/ml) |
| Penicillin | Ampicillin (AMP) (10 µg/ml) |
| Tetracyclines | Tetracycline (OXT) (30 µg/ml) |
| Sulphonamides/ Trimethoprim | Co-trimoxazole (CTX) (25 µg/ml) |

*E. coli* ATCC 25922(American Type Culture collection, Rockville, MD, USA) was used as reference control strain.

### Phenotypic Screening of *Enterobacteriaceae* for Extended-spectrum β-lactamases (ESBLs) production

Cefotaxime *(30μg)* and Ceftazidime *(30μg)* antibiotic discs were used to phenotypically test Enterobacteriaceae for the development of ESBLs and incubated overnight at 37 ^0^C on Muller Hinton agar (16).

### Confirmation of ESBL producing Enterobacteriaceae

#### Double Disc Synergy method

A sterile cotton swab was used to surface spread an Enterobacteriaceae suspension on to a Mueller Hinton agar plate after turbidity was set to the 0.5 McFarland standards. The antibiotic discs used were Ceftazidime (CAZ) (30μg) alone and Ceftazidime in combination with Clavulanic acid (CAL) (30/10μg) (17). The discs were spaced approximately 30 mm apart, and the discs and plates containing the test organism were incubated at 37° C overnight. Both the single disc and the combined disc’s zones of inhibition were measured. Clavulanic acid-containing discs with an increase in zone diameter of ≥5mm in comparison to those without Clavulanic acid were deemed to have an ESBL (17).

### Genomic DNA extraction

Bacterial genomic DNA extraction was conducted on all the 18 Salmonella and 57 Pathogenic E coli from the pure plate colonies that were extracted. The Bio line ISOLATE II genomic DNA kit (Cat No. Bio-52065 Lot No. IS502-B054750) was used for the bacterial extraction following manufacturer’s instruction. The presence of genes encoding for ESBL (*bla*-TEM gene) was detected using conventional PCR amplification using primers listed in (table 2).

**Table 2:**
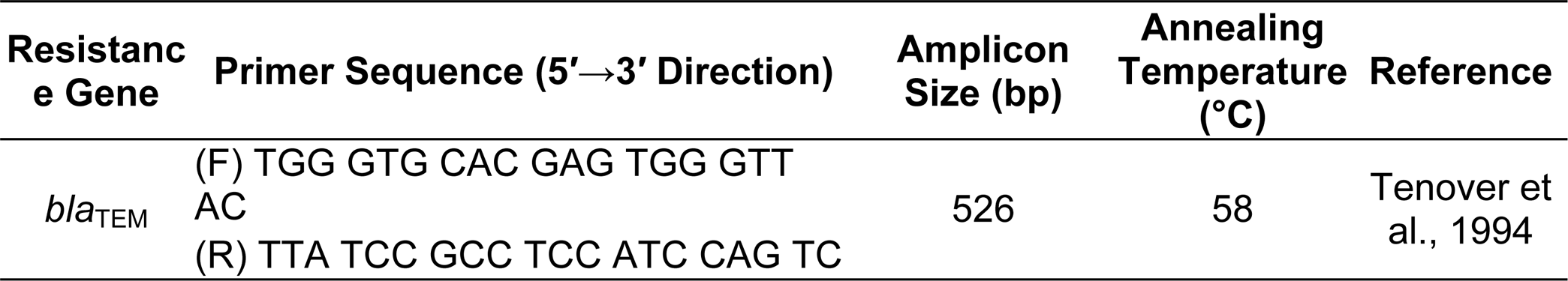
Primers used in the PCR reaction.

The PCR master Mix reagents was prepared as follows:

- 12.5 µL master mix consisting of One Taq quick load two times master mix/w standard buffer,
- dNTPs&Taq polymerase (M0486S),
- 1.5 µL forward (100 µM),
- 1.5 µL primary reverse (one hundred µM),
- 5 µL DNA template and RNAse-free dH2O up to 25 µL.

**PCR Cycling:** The PCR process was carried out in a thermo cycler (Perkin Elmer, Wellesley, MA, USA) with a pre-denaturation cycle of 95°C for 15 min, followed by DNA amplification stage with 30 cycles (94°C for 1 min, 58°C for 1 min, and 72°C for 1 min) and final extension cycle of 72°C for 5 min.

DNA Amplicons were electrophoresed using 1.5% agarose gel, in Tris-Borate EDTA buffer (TBE) 1×concentration, Safe View Classic^TM^ DNA stain, 6x loading dye (Thermo Scientific), and DNA ladder/marker 100 bp (Sigma-Aldrich, Inc., Saint Louis, MI, USA)DNA Bands were visualized on a Dark reader Transilluminator.

### Data Management and Analysis

All records of the analysis were recorded in the lab register as a hard copy back up. Samples were assigned codes and excel spreadsheets were used to enter the raw data, which were then exported to Stata (Version 12, Special Edition, College Station, Texas USA) software for statistical analysis. Frequency tables, graphs were used to present descriptive statistics. This was done at a univariate analysis level.

Bivariate and multivariate analyses was carried out for the study objectives, quantitative data evaluations was made in relation to the 95% level of significance/confidence, the Pearson value (*p-value*) to determine the objective variables’ statistical significance, statistics were judged to be significant for any p < 0.05.

### Ethical Consideration

Approval was obtained from Mbarara University of Science and Technology; Institutional Ethical Review Committee (MUST-2021-141), and at the ministry level, permanent secretary Ministry of Agriculture, Animal Industries, and Fisheries, district’s chief administrative officer, and district veterinarian.

Prior engaging the farms, the researcher sought for clearance from the farm owners to access and collect samples from their farms. This consent was requested voluntarily to participate in the study. The researcher treated all Farm’s data and bacterial isolates obtained for this investigation in strict confidence.

### COVID19 Prevention and Management Plan

The researcher received the recommended two doses of COVID-19 vaccination. In addition to this, the researcher adhered to the government recommended standard operating procedures such as avoiding touching surfaces at the poultry farms on any place while carrying out this study, frequent washing/cleaning of the hands with soap or sanitizers, strict face masking while carrying out this study and the respondents will also be encouraged to put on their face masks. Social distancing was also adhered while interacting with persons at the time of data collection.

### Quality assurance and Quality control procedure

Involved the use of reference controls (both positive and negative). During the data entry and analysis, the researcher ensured double entry of data to rule out clerical errors.

## RESULTS

### Prevalence of Salmonella and E. coli from the cultured samples

Of the total of 216 samples collected, a total of 40 (18.5%) *Salmonella* and 120 (55.6%) Pathogenic *E. coli* was isolated as seen in the flow chart (figure 2).

**Figure 2:**
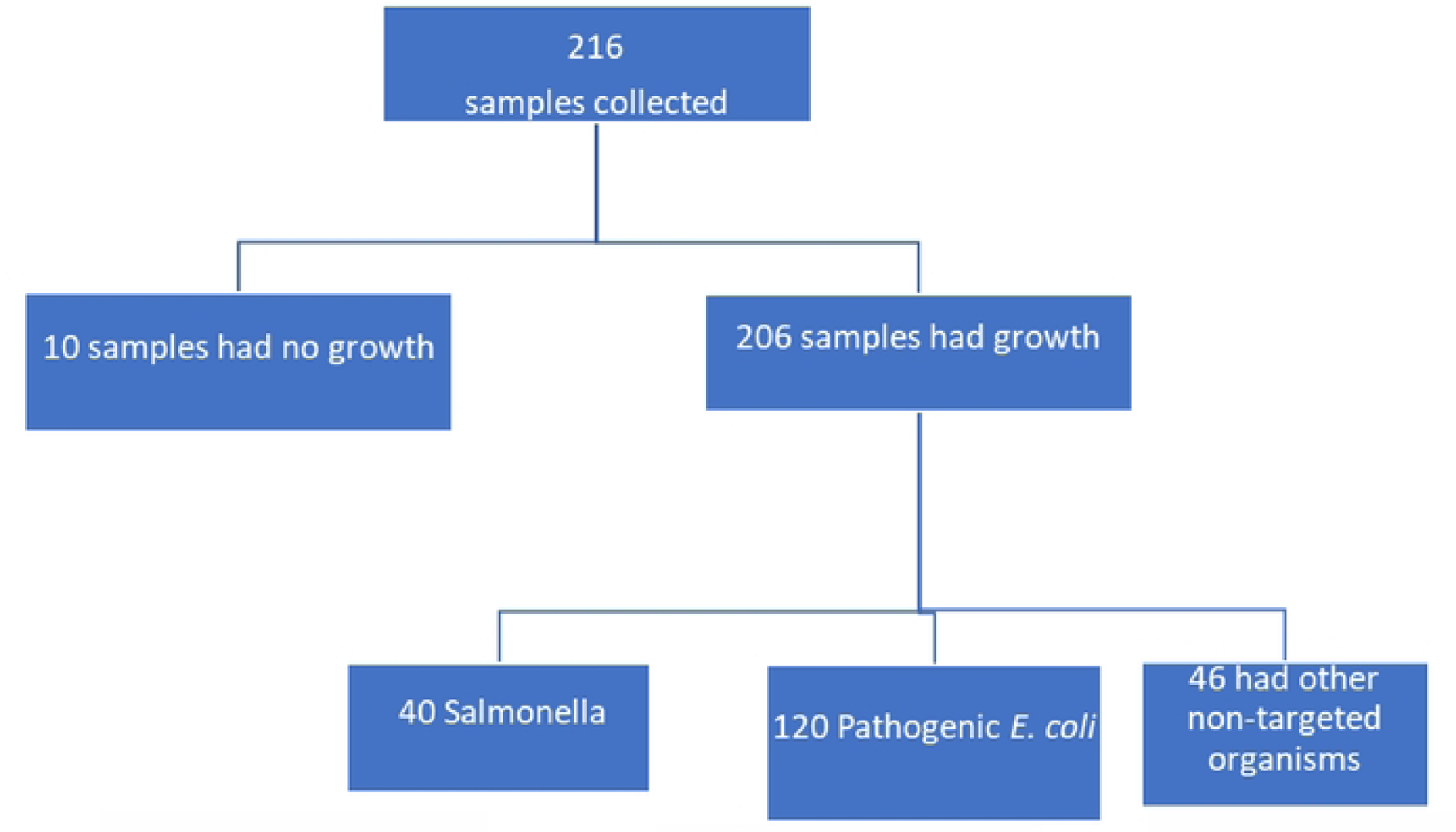
Flow chart:

### Demographic and socio-economic characteristics of the respondents

Out of 216 farm managers selected from ten (10) different sub counties in Wakiso district that took part in the research and were interviewed, 87 (40.28%) were Males and 129 (59.72%) were females. The age group of31-45years had the most participants at 113 (52.31%) while the age group of 46 and older had the fewest 50 (23.15%). In regards Education level training; at least all the farm managers had some basic training. Participants who completed primary level were 58 (26.85%), Secondary level were 80 (37.04%), vocational training were 33 (15.28%) and University level were 45 (20.83%). When examining the various farm managers’ sources of income, the survey discovered that 146 participants (67.59%) relied primarily on poultry, others had mixed agricultural practices like livestock farming (including either cattle, Sheep, Goats or Piggery) alongside poultry 22 (10.18%), crop farming with poultry 17 (5.56%) and others had additional funds arising from self-employment in other sectors alongside the poultry 31 (14.35%) as seen in table 3.

**Table 3:** Farm characteristics and demographics:

| Characteristics<br>(Variable) | Category | Frequency<br>(N=216) | Percentage<br>(%) |
| --- | --- | --- | --- |
| Gender | Male | 87 | 40.28 |
|  | Female | 129 | 59.72 |
| Age(years) | 15-30 | 53 | 24.54 |
|  | 31-45 | 113 | 52.31 |
|  | >=46 | 50 | 23.15 |
| Education level | Primary | 58 | 26.85 |
|  | Secondary | 80 | 37.04 |
|  | Vocational Training | 33 | 15.28 |
|  | University | 45 | 20.83 |
| **Sub counties** | Kitabi | 25 | 11.57 |
|  | Namugongo | 28 | 12.96 |
|  | Kiira | 32 | 14.81 |
|  | Makindye | 28 | 12.96 |
|  | Nabweru | 19 | 8.79 |
|  | Kakiri | 21 | 9.72 |
|  | Busukuma | 11 | 5.09 |
|  | Wakiso TC | 16 | 7.40 |
|  | Masulita | 12 | 5.55 |
|  | Nsangi | 24 | 11.11 |
| **Source of income** | Poultry alone | 146 | 67.59 |
|  | Livestock farming (Cattle, goats, sheep, or Pigs) with poultry | 22 | 10.18 |
|  | Crop farming with Poultry | 17 | 5.56 |
|  | Self-employed off-farm. (Others) | 31 | 14.35 |
| **hygiene/rate of cleaning of the farm** | Once per Month | 13 | 6.02 |
|  | Once per week | 117 | 54.17 |
|  | Daily | 86 | 39.81 |
| **Presence of other animals (Pigs, Cows, Ducks and Turkey)** | Other Birds (Ducks, Turkey, and Geese) | 68 | 31.48 |
|  | Livestock | 148 | 68.52 |
| **Type of production system used** | Deep Litter | 103 | 47.69 |
|  | Battery Cage system | 82 | 37.96 |
|  | mixed farming with turkey and ducks | 19 | 8.76 |
|  | Semi intensive (Free range system) | 12 | 5.56 |

### Factors associated with Salmonella and Pathogenic E. coli in broiler poultry farms in Wakiso District

*Salmonella* and pathogenic *E. coli* were most frequently found in broiler poultry farms where there was contact between poultry and other bird species like turkeys and geese (*P-value*= 0.019), contamination of commercial poultry feeds (*P*-value=0.012), movement of farm workers between pens (*P*-value=0.167)and use of untreated water (*P*-value=0.117). Therefore, there is a significant association between the above variables and the presence of the organisms of interest in the research (table 4).

**Table 4:** Showing factors associated with Salmonella and E. coli in poultry farms in Wakiso District.

| Variable | Category | Salmonella | P-value | E. coli | P-value |
| --- | --- | --- | --- | --- | --- |
|  |  | Frequency (n)=40 |  | Frequency (n)=120 |  |
| Contact of poultry and other bird species | Livestock | 83 (69.2) | 0.384 | 65 (81.25%) | 0.020 |
|  | Other birds | 25 (20.8) |  | 15 (18.75%) |  |
| Frequency of cleaning and disinfection of the Bird's Housing. | Daily | 96 (80) | 0.498 | 49 (61) | 0.113 |
|  | Once a week | 21 (17) |  | 26 (33) |  |
|  | Once a Month | 03 (3) |  | 05 (6) |  |
| contamination of commercial poultry | Mixed with Water | 95 (79.2) | 0.242 | 56 (70) | 0.002 |
|  | Mixed with other materials | 25 (20.8) |  | 24 (30) |  |
| movement from one pen to the other by farm-handlers | From one pen | 58 (48.3) | 0.244 | 55 (68.75%) | 0.017 |
|  | From more than one pen | 62 (51.7) |  | 25 (31.25%) |  |
| use of untreated water | Use of well water | 38 (31.7) | 0.392 | 32 (40) | 0.018 |
|  | Use of tap water | 82 (68.3) |  | 48 (60) |  |
| Source of poultry feeds | Home made | 46 (38.3) | 0.066 | 25 (31) | 0.237 |
|  | Commercial | 74 (61.7) |  | 55 (69) |  |

### Antibiotic susceptibility patterns of Salmonella and Pathogenic E. coli towards commonly used antimicrobials in poultry

The phenotypic resistance profile for Salmonella organisms from broiler poultry samples when subjected to the selected antibiotics demonstrates that ampicillin 32 (80%) is the drug with the highest level of resistance, followed by erythromycin 28 (70%), tetracycline 27 (67.5%), and ciprofloxacin 25 (62.5%). On the contrary, low resistance was observed to ceftazidime 10 (15%), Gentamicin 3 (8%), Chloramphenicol 3 (8%), cefepime 2 (5%) and meropenem2 (5.0%) as shown in table 5.

**Table 5:** showing antibiotic susceptibility patterns of Salmonella towards commonly used antimicrobials in Poultry:

| Antibiotics | Resistant<br>n (%) | Intermediate<br>Resistance n<br>(%) | Susceptible<br>n (%) |
| --- | --- | --- | --- |
| Gentamicin (GM) (10 µg/ml) | 3 (7.5) | 7 (17.5) | 30 (75.0) |
| Meropenem (MEM) (10 µg/ml) | 1 (2.5) | 2 (5.0) | 37 (92.5) |
| Ceftriaxone (CRO) (30 µg/ml) | 18(45) | 4 (10) | 18 (45) |
| Chloramphenicol (CHL)<br>(30 µg/ml) | 3 (7.5) | 3 (7.5) | 34 (85) |
| Cefepime (CPM) (30 µg/ml) | 2 (5.0) | 3 (7.5) | 35 (87.5) |
| Ciprofloxacin (CIP) (5 µg/ml) | 25 (62.5) | 10 (25) | 5 (12.0%) |
| Erythromycin (EM) (30 µg/ml) | 28 (70) | 7 (17.5) | 5 (12.5) |
| Ampicillin (AMP) (10 µg/ml) | 32 (80) | 2 (5) | 6 (15) |
| Tetracycline (OXT) (30 µg/ml) | 27 (67.5) | 6 (15) | 7 (17.5) |
| Sulfamethoxazole-trimethoprim<br>(SXT) (25 µg/ml) | 20 (50) | 5 (12.5) | 15 (37.5) |
| Cefotaxime (CTX)(30µg/ml) | 10 (25) | 2 (5) | 28(70) |
| Ceftazidime (CTZ) (30µg/ml) | 15 (37.5) | 5 (12.5) | 20 (50) |

E. coli exhibited the highest resistance to Erythromycin at 88 (73%), followed by Ampicillin 86 (72%), Chloramphenicol 85 (71%), Tetracycline at 82 (68%) and sulfamethoxazole-trimethoprim at 78 (65%). However, it’s worth noting that there was observed very low resistance to Ciprofloxacin 9 (8%), meropenem 4 (3%), and cefipime 3 (2.5%) as seen in table 6.

**Table 6:**
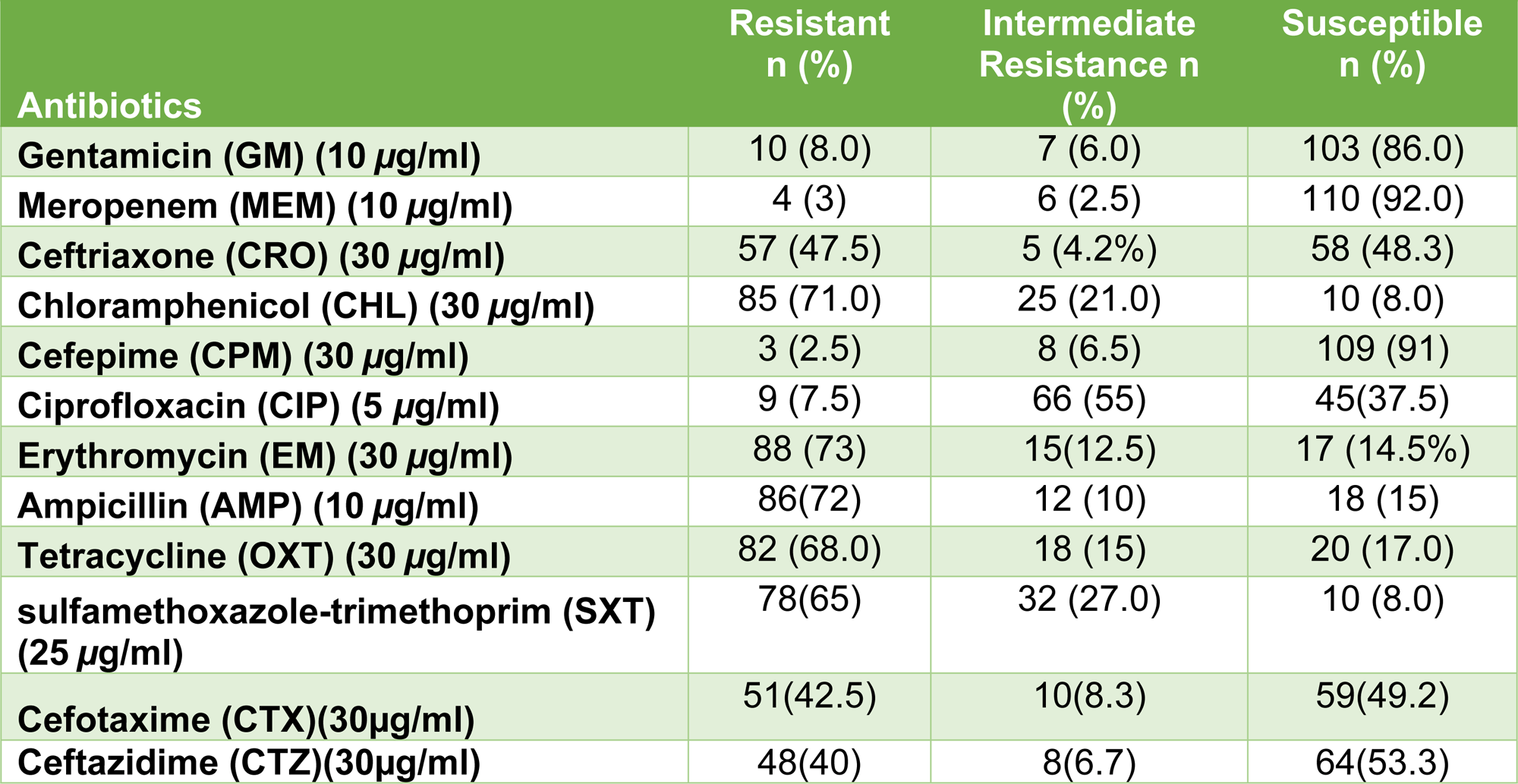
showing antibiotic susceptibility patterns of pathogenic *E. coli* Spp towards commonly used antimicrobials in Poultry.

| Antibiotics | Resistant<br>n (%) | Intermediate<br>Resistance n<br>(%) | Susceptible<br>n (%) |
| --- | --- | --- | --- |
| Gentamicin (GM) (10 µg/ml) | 10 (8.0) | 7 (6.0) | 103 (86.0) |
| Meropenem (MEM) (10 µg/ml) | 4 (3) | 6 (2.5) | 110 (92.0) |
| Ceftriaxone (CRO) (30 µg/ml) | 57 (47.5) | 5 (4.2%) | 58 (48.3) |
| Chloramphenicol (CHL) (30 µg/ml) | 85 (71.0) | 25 (21.0) | 10 (8.0) |
| Cefepime (CPM) (30 µg/ml) | 3 (2.5) | 8 (6.5) | 109 (91) |
| Ciprofloxacin (CIP) (5 µg/ml) | 9 (7.5) | 66 (55) | 45(37.5) |
| Erythromycin (EM) (30 µg/ml) | 88 (73) | 15(12.5) | 17 (14.5%) |
| Ampicillin (AMP) (10 µg/ml) | 86(72) | 12 (10) | 18 (15) |
| Tetracycline (OXT) (30 µg/ml) | 82 (68.0) | 18 (15) | 20 (17.0) |
| sulfamethoxazole-trimethoprim (SXT)<br>(25 µg/ml) | 78(65) | 32 (27.0) | 10 (8.0) |
| Cefotaxime (CTX)(30µg/ml) | 51(42.5) | 10(8.3) | 59(49.2) |
| Ceftazidime (CTZ)(30µg/ml) | 48(40) | 8(6.7) | 64(53.3) |

### Detection of ESBL (*blaTEM*), gene encoding resistance to commonly used antibiotics used in poultry in Salmonella and Pathogenic E. coli

Out of the 18 *Salmonella* samples analyzed for genotypic expression, 7/18 (39%) samples expressed presence of the *bla* TEM genes while 11 (61.1%) samples did not have this gene.

Out of the 57 Pathogenic *E. coli* samples analyzed for genotypic expression, 42/57 (73.8%) samples expressed presence of the *bla TEM* genes while fifteen (26.3%) samples did not have this gene.

## DISCUSSION

### The prevalence of selected Salmonella and pathogenic E. coli in selected poultry farms

According to this study, a prevalence of 18% and 56%, respectively, was identified for *Salmonella* and pathogenic *E coli* in broiler poultry farms in the Wakiso district. In line with other studies carried out, this result was comparable to a study conducted by (18). This revealed the prevalence of *Salmonella* and pathogenic *E. coli* as 21.1% and 56.3% in Uganda.

However, it can be argued that this prevalence rate is low when compared to a study done by (19) found an overall prevalence of 83%, of which 90.8% and 73% were from chicken in Lira and Kampala districts from the antibiotic susceptibility profiles of fecal Escherichia coli isolates from Dip-Litter broiler chicken in Northern and Central Uganda.

This discrepancy can be due to the different study sites, sample methods, poultry sector, and sampling times used during the research.

It is worth noting that this study had a lower prevalence of *Salmonella* spp compared to other studies such as one done by (20) in Ruiru Sub-County, Kenya which was at 28% with almost similar prevalence of pathogenic *E. coli* of 58%. Another study on antimicrobial resistance in *Salmonella* and *Escherichia coli* isolates from chicken droppings in Nairobi (21) found lower levels of Salmonella (12% vs. 57% in our study) and nearly similar prevalence of E. coli (57%) in the analyzed samples.

The differences in environmental contamination levels, poultry management practices, breed, sample size, sampling, testing methodologies, and challenges in *Salmonella* detection methods may account for this similar trend of reduced *Salmonella* isolation and prevalence (22) or further still the practice of better bio safety and bio security practices at farms overseen by the established of Kenya Accreditation society (KENAS),competitive exclusion of sick birds, breeding for genetic resistance and vaccination.

In Uganda as evidenced from the essential veterinary medicines list, vaccines exist only in private practitioners’ clinics and cannot be accessed freely by the Bio security level three farmers that were of interest in this study.

### The factors associated with Salmonella and Pathogenic E. coli in poultry farms in Wakiso District

Presence of *Salmonella* and pathogenic *E*. *coli* in the poultry farms was significantly correlated with contact of poultry with other avian bird species such as ducks, geese, and guinea fowls in the same farms and pens, as well as not frequently cleaning the poultry farms by removing the manure or beddings.

The study further investigated other factors associated with proper poultry practices such as the implementation of strict bio security interventions such as having restricted access to the farmers by visitors and handling of these birds, implementation of a solid well established sewer system. These, however, did not significantly increase the risk of *Salmonella* and *E. coli*. This is comparable to a study conducted in Nigeria that focused on the risk factors related to *Salmonella* spp. In broiler and layer flocks, it was discovered that the presence of rodents, farm workers moving between pens, running and parking trucks close to poultry farms (p<0.05) and drinking untreated water (p<0.05) were all independently associated with a higher risk of *Salmonella* infection (23).

Broilers are known for consuming large amounts of feed, and this habit encourages constant feces loss, raising the possibility of their environment becoming contaminated with various bacterial strains (18).

Furthermore, many broiler farms had high stock densities, which could make environmental management efforts to reduce bacteria in the houses more difficult. As a result, workers (especially those handling large flocks) must be strictly supervised because it is claimed that they may neglect their responsibilities for maintaining hygiene. Therefore, poor management of poultry could lead to increased transmission of *Salmonella* and *E. coli* in poultry.

### The antimicrobial Susceptibility patterns of Salmonella and E. coli in poultry farms

The highest resistance to ampicillin was found in both *Salmonella* and *E. coli* isolates, 32 (80%) and 86(72%), followed by erythromycin (28%) and 88(73%) and tetracycline (27) and 82(68.0%).

Our research was comparable to a study published by (24) and (18). Therefore, there is great increase in antimicrobial resistance to the different drugs most especially ciprofloxacin in Uganda.

According to reports from Uganda and other nations (25), tetracyclines are frequently used to treat bacterial illnesses and promote animal growth (26). Therefore, it is not surprising that pathogens have developed broad resistance to them. Bacteria like commensal E. coli experience selection pressure as a result of ongoing exposure to antimicrobials (27).

In most nations, the rise in human cases of antimicrobial resistance is attributed to the increase and spread of infections from poultry to humans. In this work, we discovered that pathogenic *E. coli* and *Salmonella* spp. isolated from chicken in the Wakiso district had high resistance to widely used antibiotics used in both people and animals. As a result, greater research on AMR in *Salmonella* spp. and *E. coli* clinical isolates from poultry is needed.

### ESBL producing genes present among Salmonella and pathogenic E. coli isolated from poultry farms

The majority of studies on poultry have noted the presence of genes such as; *bla*TEM, AmpC like lactamase gene, for antibiotic resistance (28), (29) and (30). A study conducted in Malaysia reported a lower prevalence of *bla*TEM (31) while a study in British Columbia, 2007 reported slightly higher levels of the *bla*TEM gene, being 81.5 and 80% of amoxicillin and ampicillin resistant *E.coli* and *Salmonella* spp. isolates (32).

Therefore, the community’s health is at risk because of the possibility that antimicrobial resistance genes in poultry waste will spread to humans and other fowl.

## CONCLUSION

The two most significant food-borne pathogens of public health concern linked to poultry are still *Salmonella* spp. and *Escherichia coli*, and it is evident from this study with a prevalence of 18% and 56% respectively. These bacteria have resistant genes associated with them seen in 38.9% and 73.8% *Salmonella* and pathogenic *E. coli* samples.

### Limitations

i. Our focus in this study was strictly broiler poultry that were about to enter the food chain, this is does not paint a full picture in terms of the overall burden of the disease in poultry including all other avian birds such as layers, geese, guinea fowls and ducks to effectively understand the disease transmission dynamics and effect policies that will effectively halt further spread.
ii. Only one gene blaTEM was focused on during this study and it would be vital to conduct different genetic manipulations of the same bacterial DNA to targeting different ESBL genes such as bla CTM, bla SHV etc.

### Recommendations

Based on the above the study findings, its pertinent that government of Uganda should strengthen the antimicrobial stewardship program and ensure that it is supported to carry out its mandate of coordination that supports the proper use of antimicrobials (including antibiotics), improves patient outcomes, lowers microbial resistance, and limits the spread of diseases brought on by multidrug-resistant organisms, the organization needs help.

This should be achieved through the one health platform, a forum that brings together all key stakeholders from the four different line ministries of Health, Ministry of Agriculture Animal Industries and Fisheries, Ministry of Water and sanitation and Ministry of tourism, trade, and antiquities.

## Data Availability

The authors confirm no conflicts of interest pertaining the publication of this article.

N.A

## DECLARATION

### Author contributions

TS and KT developed the study concept, TS, JCB and NPP provided input in data collection. TS and JCB analysed the data. KT and JB over saw the entire study. NPP, KT and JB wrote the final manuscript. All authors read and commented on the paper and agreed on the final version.

### Availability of data and materials

The analyzed datasets are available from the corresponding author upon request.

### Consent for publication

Not applicable

